# Mapping open educational resources on how to justify, design, conduct, analyse, and share randomised clinical trials: a landscape analysis

**DOI:** 10.1101/2025.05.16.25327604

**Authors:** Kim Boesen, Frances Shiely, Shaun Treweek, Sarah Louise Klingenberg, Christian Gluud

**Affiliations:** Copenhagen Trial Unit, Centre for Clinical Intervention Research, The Capital Region, Copenhagen University Hospital ─ Rigshospitalet, Copenhagen, Denmark; HRB Clinical Research Facility, University College Cork, Cork, Ireland; School of Public Health, University College Cork, Ireland; Aberdeen Centre for Evaluation, University of Aberdeen, Aberdeen, Scotland; Department of Regional Health Research, The Faculty of Health Sciences, University of Southern Denmark, Odense, Denmark

**Author notes:** **Corresponding author** Kim Boesen, MD PhD.

## Abstract

**Objective:** To map open educational resources on how to justify, design, conduct, analyse, and share randomised clinical trials of healthcare interventions.

**Design:** Landscape analysis.

**Data sources:** Systematic searches of multiple databases of biomedical literature using ASReview, a machine learning assisted systematic review tool, to screen the most relevant records. Manual searches of general websites, regulatory and clinical trial unit websites, and online learning portals.

**Main outcomes:** Categorisation and reporting of included resources according to type (e-learning, dedicated trial platform, scattered website material, e-books, and videos), format (guidance, templates, research publications, and tools), and trial stage content (justification, design, conduct, analysis, and sharing).

**Results:** We included 63 open educational resources categorised as dedicated trial portals (n=22), e-learning courses (n=20), scattered website material (n=10), videos and webinars (n=9), and e-books (n=2). Various formats were used and the content distribution according to trial stage was skewed towards design (n=47) and conduct (n=51), with fewer resources on justification (n=12), analysis (n=27), and sharing (n=23). Few resources provided clear and instructive navigation roadmaps for users or catalogued their content according to the clinical trial lifecycle.

**Conclusion:** We identified a range of open educational resources on how to justify, design, conduct, analyse, and share randomised clinical trials. Most content focused on trial design and conduct, whereas some aspects of trial justification were entirely absent. Most resources lacked a clear roadmap to catalogue the content. The available resources provide valuable and extensive information, but users must be wary of important gaps in the existing landscape of educational material on randomised clinical trials. New open educational resources should provide clear roadmaps and address the underprioritised areas of the trial life cycle.

**Review registration:** https://doi.org/10.1101/2024.02.16.24302873 (preprinted 17 February 2024).

**Summary boxes:** *What is already known on this topic:*

- Most conducted randomised clinical trials do not inform clinical practice and may be considered research waste.
- A lack of access to learning materials on how to justify, design, conduct, analyse, and share randomised clinical trials may be one possible factor to research waste.
- We decided to map open educational resources on randomised clinical trials to create a one-stop collection and to identify gaps in the landscape.

*What this study adds:*

- We identified an array of open educational resources, covering large parts of the clinical trial life cycle.
- There are important gaps in the landscape, most importantly regarding the justification of new randomised clinical trials.

## Introduction

Many randomised clinical trials (RCTs) of healthcare interventions do not inform clinical practice and are considered research waste.^1–7^ Trials may not be informative if they are not justified by systematically considering previous research, not designed optimally, incorrectly analysed, and not reported transparently.^8, 9^ Recent analyses of clinical trials’ informativeness assessed various indicators such as scope (i.e. was the clinical question relevant), feasibility (i.e. was the trial possible to conduct), reliability (i.e. were the trials at risk of bias), and transparency (i.e. were the methods and results completely reported). One study of 125 diabetes, ischaemic heart disease, and lung cancer trials found only 26% informative.^10^ Another study of 347 gynaecology and obstetrics trials found only 10% of trials informative.^11^ Meta-epidemiological analyses corroborate these assessments; nine of 10 trials included in recent Cochrane systematic reviews were rated as high or unclear risk of bias,^12^ and only one in 10 effective healthcare interventions were supported by high quality evidence.^13^

There may be several reasons for this lack of ‘informativeness’ in contemporary clinical trials: it may not be a priority during graduate and postgraduate training to teach the scientific reasoning behind clinical trials; access to freely available quality educational material may be sparse; and researcher incentives focus on prestigious academic publications and grant funding, not on generating actionable evidence. Finally, current clinical trial regulations do not focus on informativeness but rather on aligning with International Council of Harmonisation (ICH) - Good Clinical Practice (GCP) standards.^14, 15^ These GCP standards are criticised for making trials overly bureaucratic^16^ and their introduction was associated with a reduction in the number of trials conducted in Europe.^17^ It is unknown if GCP improves informativeness and reduces research waste.

Many commercial providers offer clinical research training to pharmaceutical companies and clinical research organisations, which focuses primarily on GCP.^18^ To facilitate training for both industry and academic trial teams and ensure the work invested in developing existing materials is not wasted, we wanted to map publicly available online educational material covering the whole life cycle of RCTs of healthcare interventions; from the initial ideation and justification to the design, conduct, analysis, and final sharing of results and data. We focused on free online material, colloquially known as open educational resources (OER),^19, 20^ which is often used in a ‘blended learning’ setup combining face-to-face classes with online self-study.^21, 22^

## Methods

We wished to identify publicly available educational material or open educational resources on the justification, design, conduct, analysis, and sharing of RCTs. We report our analysis according to the PRISMA reporting guideline for scoping reviews.^23^ Our protocol was published 17 February 2024.^24^

### Eligibility criteria

We defined open educational resources as any “learning, teaching and research material in any format and medium that reside in the public domain”^20^ on how to justify, design, conduct, analyse, and share RCTs. We searched for any free, publicly available, e-learning courses or other educational resources and platforms covering any aspect of the clinical trial life cycle. We included resources that we deemed had potential to teach those interested in conducting RCTs. Generic information about RCTs targeted mainly at the public, e.g. Wikipedia, were not included.

We applied no restrictions on language, publication year, and content, nor did we define a cut-off based on the granularity and level of detail. Content that required registration was also included, provided anybody could access it. We did not search for or include solitary research publications, e.g. guidance or instructions on RCTs published in traditional medical journals, for two reasons; it would require searching systematically on each of the five trial stages, which would constitute a very extensive task, and many research papers are behind paywalls and thus would not be eligible for inclusion. Instead, we opted to search databases of published literature for papers describing OER or other comparable initiatives (e.g. websites or online courses) eligible for inclusion.

### Search strategies

We used a two-step approach by first ‘cold searching’ general websites using expected key words to identify ‘seed references’ such as relevant material, videos, and resources. One author (KB) used these keywords and terminology to subsequently search the listed resources. We report exhaustively each searched resource and search string in the Datafile (see link in data sharing statement).

#### Systematic search of medical literature databases

To identify papers describing potentially relevant resources, the information specialist (SLK) searched MEDLINE Ovid, Embase Ovid, Web of Science, and CINAHL (EBSCO) using a broad scoped compound search string consisting of keywords relevant to the field. See Appendix, Supplement 1 for details.

#### General websites

We searched general websites to find relevant course material, including YouTube, Google, and Bing, screening the first 100 hits for each search.

#### Online e-learning portals

We searched ClassCentral, Coursera, and EdX. We did not search paid-only portals, such as Udemy, the Elsevier-owned platform Osmosis, FutureLearn, Udacity, or other learning platforms like Online MedEd, which is a clinically oriented platform.

#### University online learning portals

We searched a list of US and European university online learning portals.

#### Clinical trial units

We searched clinical trial unit websites from Denmark, Sweden, Norway, Switzerland, United Kingdom, United States, and international clinical trial networks.

#### Clinical trial registries and national authorities

We searched the websites of clinical trial registries and national regulatory authorities.

### AI assisted screening

We used the open-source, machine learning assisted screening tool for systematic reviews, ASReview^25^ to screen the obtained records from our systematic search. One author (KB) screened the hits. The included hits were subsequently assessed in detail for a final decision, and in case of uncertainty another author (CG) was consulted to reach a decision. We had defined an arbitrary stopping rule of 100 consecutive irrelevant hits in our protocol, but we decided to expand the screening to 1% of the total sample. We trained the ASReview screening algorithm on four relevant and five irrelevant records (Appendix, Supplement 2).

### Categorisation of OERs

#### Classification by type

We classified the identified resources into five different types: (1) **e-learning courses** (e.g. various material presented as video, text, or exercises followed by tests and a certification of completion); (2) **dedicated trial portals** (e.g. websites dedicated for learning purposes on RCTs); (3) **scattered website material** (e.g. material found on websites dedicated to other non-learning purposes); (4) **e-books** (e.g. extensive manuals or full-stack books); and (5) **videos and webinars** (i.e. either solitary or collections).

#### Classification by formats

We categorised content formats as: (1) **guidance/tutorial/instructions** (e.g. material intended to teach or give instructions on a certain matter either as video or written material); (2) **research publications** (e.g. research papers usually describing empirical work or collections of links to publications); (3) **documentation templates** (e.g. checklists or legal documents, such as Standard of Procedures or regulatory applications); (4) **tools** (e.g. applications to help design, conduct, or analyse clinical trials, e.g. sample size calculator or randomisation tools). Each resource could contain multiple formats.

#### Classification by trial stage

Finally, we categorised each OER’s content into five trial stages, depending on where it belongs in the RCT life cycle: (1) **justification** (e.g. how to search and assess the literature to decide whether a new trial is justified); (2) **design** (e.g. how to choose population, comparator, and design of other trial characteristics); (3) **conduct** (e.g. how to run the trial, adhering to GCP, and managing documentation); (4) **analysis** (e.g. how to statistically analyse outcome data after trial conduct); (5) **sharing** (e.g. how to report trial results and data in trial registries and share full datasets), table 1. We identified whether the resources had clear roadmaps mirroring the clinical trial lifecycle to navigate users.

### Patients and Public involvement

We did not involve patients or the public in this landscape analysis.

### Changes to our protocol

See Appendix, Supplement 3, for a list of all amendments compared to our published protocol.

## Results

### General search results

We identified a total of 63 OERs on how to justify, design, conduct, analyse, and share RCTs. The 23 most comprehensive resources are listed in Table 2, and all OERs are listed in the Appendix, Supplement 4. These resources were identified from our systematic search of databases (n=9), Coursera (n=9), general website searches (n=8), clinical trial unit websites (n=8), regulator websites (n=7), EdX (n=2), ClassCentral (n=1), trial registries (n=1), and from other sources (n=18). See Appendix, Supplement 4, Tables 1 to 5 for a short synopsis of each identified resource.

Our systematic search returned 196,533 records. We screened 2002 (1%) records and included 63 records for further assessment, leaving 194,531 records unscreened (Flowchart and screening recall curve, Panel 3). For details about the other searched resources, please refer to the Data File (https://doi.org/10.5281/zenodo.15410961).

**Panel 1.** Randomised clinical trial stage classification.

| Trial stage | Description |
| --- | --- |
| Justification | Description of the scientific considerations prior to deciding to conduct a randomised clinical trial. This includes the conduct of systematic reviews; deciding whether a new clinical trial is needed; and informing the design of a new trial based on previous evidence. |
| Design | How to choose the trial characteristics, such as population, intervention, comparator, or outcomes, duration and estimating the sample size. It also includes to mitigate methodological and practical limitations. |
| Conduct | How to organise and launch the trial, including coordinating stakeholder arrangements with collaborators and regulatory authorities, assembling various trial committees, adhering to Good Clinical Practice, and capture data. |
| Analysis | How to statistically analyse the various data during and after the trial conduct, including binary, ordinal and continuous outcomes, aggregate and individual patient data, and interim versus final analyses. |
| Sharing | How to prepare and report the trial results in trial registries, repositories, and in journal publications. How to share trial datasets for secondary use, e.g. for use in systematic reviews, as aggregate and individual patient data anonymised or pseudo-anonymised. |

**Panel 2.**
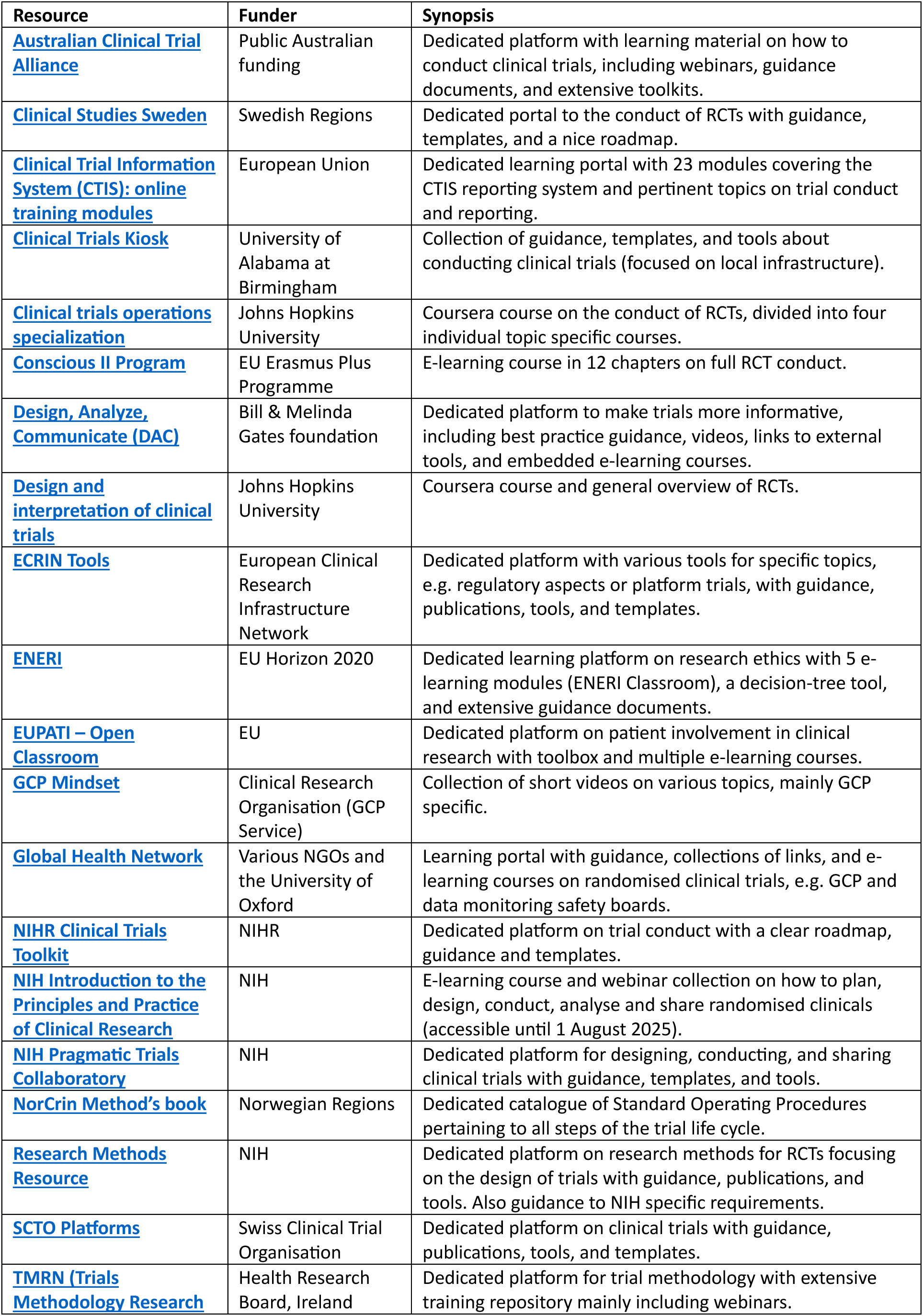
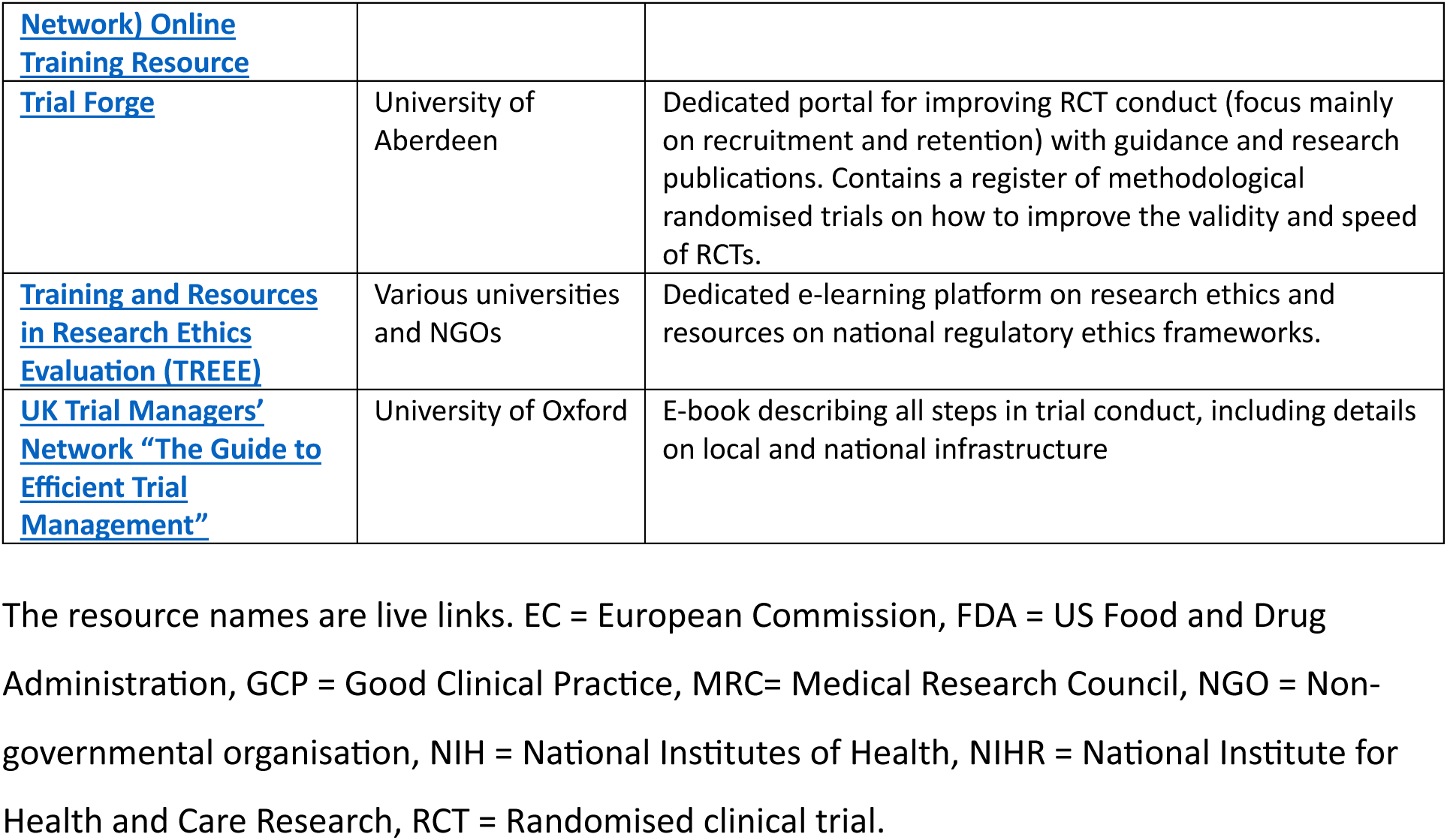
Characteristics of 23 key open educational resources.

**Panel 3.**
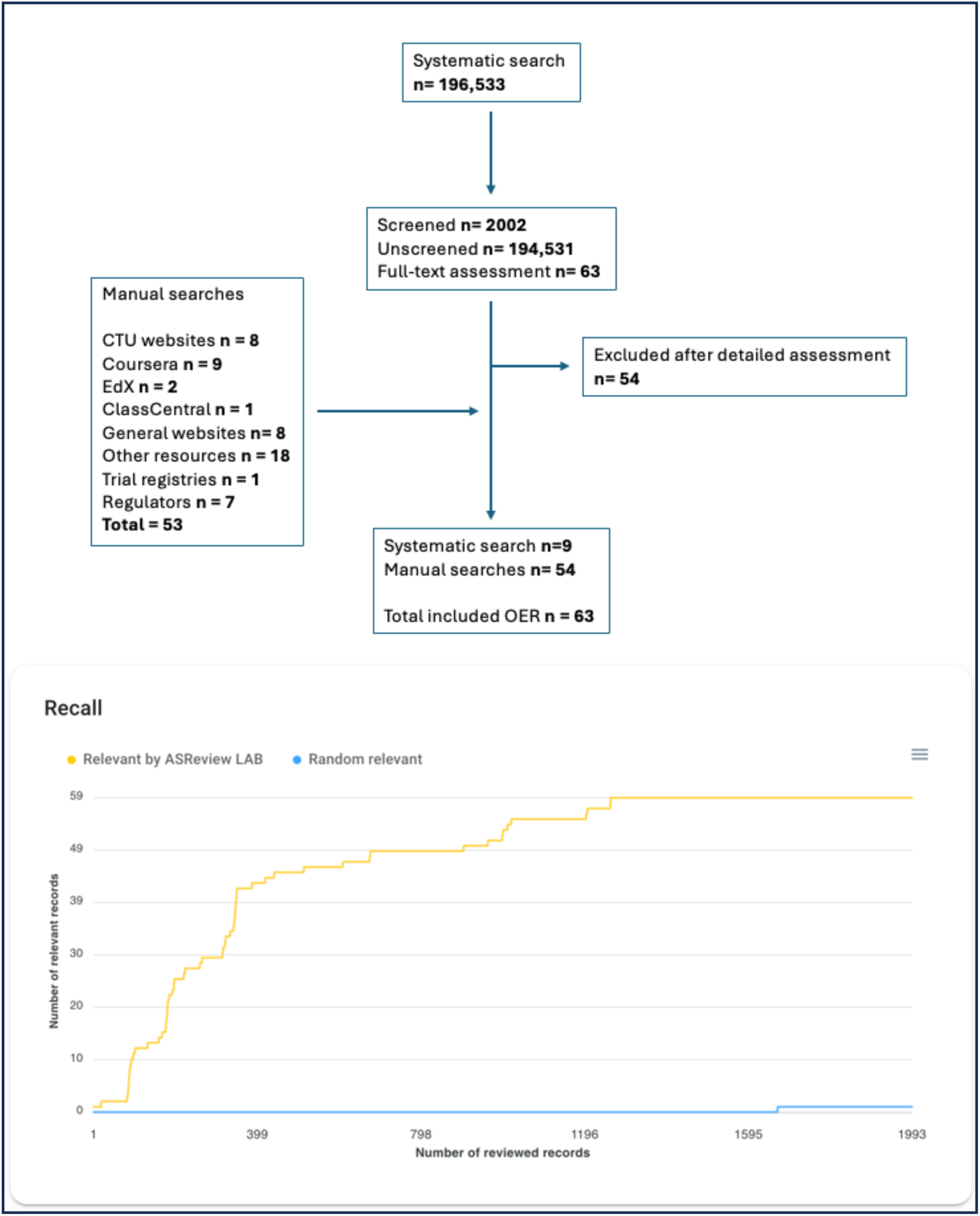
Modified PRISMA flowchart and ASReview recall curve. The yellow recall curve shows when the included hits were identified during the screening phase in ASReview. Note that the recall curve only shows the screened hits, and not the remaining unscreened hits. The bottom blue line shows a hypothetical recall curve, if the records were screened in random order.

### Classification of the identified OERs

The 63 included OERs varied enormously in scope, format, and target audience. From high-level introductory e-learning courses on clinical trials to specific courses on GCP, and from randomly scattered material on clinical trial unit websites to dedicated learning platforms.

The content varied from guidance and instructional documents, collections of links, videos, research publications, tools, and documentation templates, such as case report forms.

We classified the identified resources into five different categories: dedicated trial portals (n=22); e-learning courses (n= 20); scattered website material (n=10); videos and webinars (n=9); and e-books (n=2) (Appendix, Supplement 4, Tables 1 to 5).

### 1. Dedicated trial portals (n=22)

These were the most elaborate and extensive resources, albeit with very different scopes and content. These websites were mostly developed by large research institutions, like the National Institutes of Health (NIH), National Institute for Health and Care Research (NIHR), or the Swiss Clinical Trial Unit (SCTO) Network’s Platform. Each of these portals offered multiple resources, including guidance, instructions, tools, and template documents. Only few platforms, such as The Swedish Clinical Studies, Trial Forge, NIHR Clinical Trials Toolkit, and the NIH Pragmatic Trials Collaboratory, catalogued their content according to an explicit roadmap following the trial lifecycle. Most others provided information in a less structured fashion.

### 2. e-learning courses (n=20)

We found courses on general overview and introductions to RCTs (such as the CONSCIOUS II course) and on more specific content such as data analysis in R or how to make clinical study reports. Non-profit organisations, universities, and private companies all made e-learning courses. We identified four courses on GCP. Most courses had limited background material and references.

### 3. Scattered website material (n=10)

We identified trial unit and specific disease trial network websites that contained both general information about clinical trials and specific material on how to conduct and implement clinical trials. Some of these websites focused especially on documentation templates and specifics pertaining to local infrastructure or the disease topic. The European Medicines Agency (EMA) and FDA also publish guidance aimed primarily to inform the design and conduct of commercial trials.

### 4. Videos (n=9)

We located several video collections on clinical trials, e.g. the recorded lectures from the US Food and Drug Administration (FDA) annual clinical investigator course. One Clinical Research Organisation maintains an active YouTube channel featuring over 400 small videos, primarily on GCP topics. We also found several solitary videos on specific trial issues.

### 5. e-books (n=2)

We identified two full e-books, one describing the full clinical trial lifecycle and one specifically on data monitoring safety boards.

### Content analysis

We categorised the resource content as trial justification (n=12), design (n=47), conduct (n= 51), analysis (n=27), and sharing (n=23) (Panel 4). Individual resource content varied from high-level overviews that covered all five trial stages to narrower topic-specific resources that addressed specific aspects in detail, such as e-courses on how to compile a clinical study report. Some resources contained hundreds or thousands of individual items, guidance and links, and other resources consisted of just one document or video; thus, there is an intrinsic unit-of-analysis fallacy and the numbers should not be interpreted strictly.

**Panel 4.**
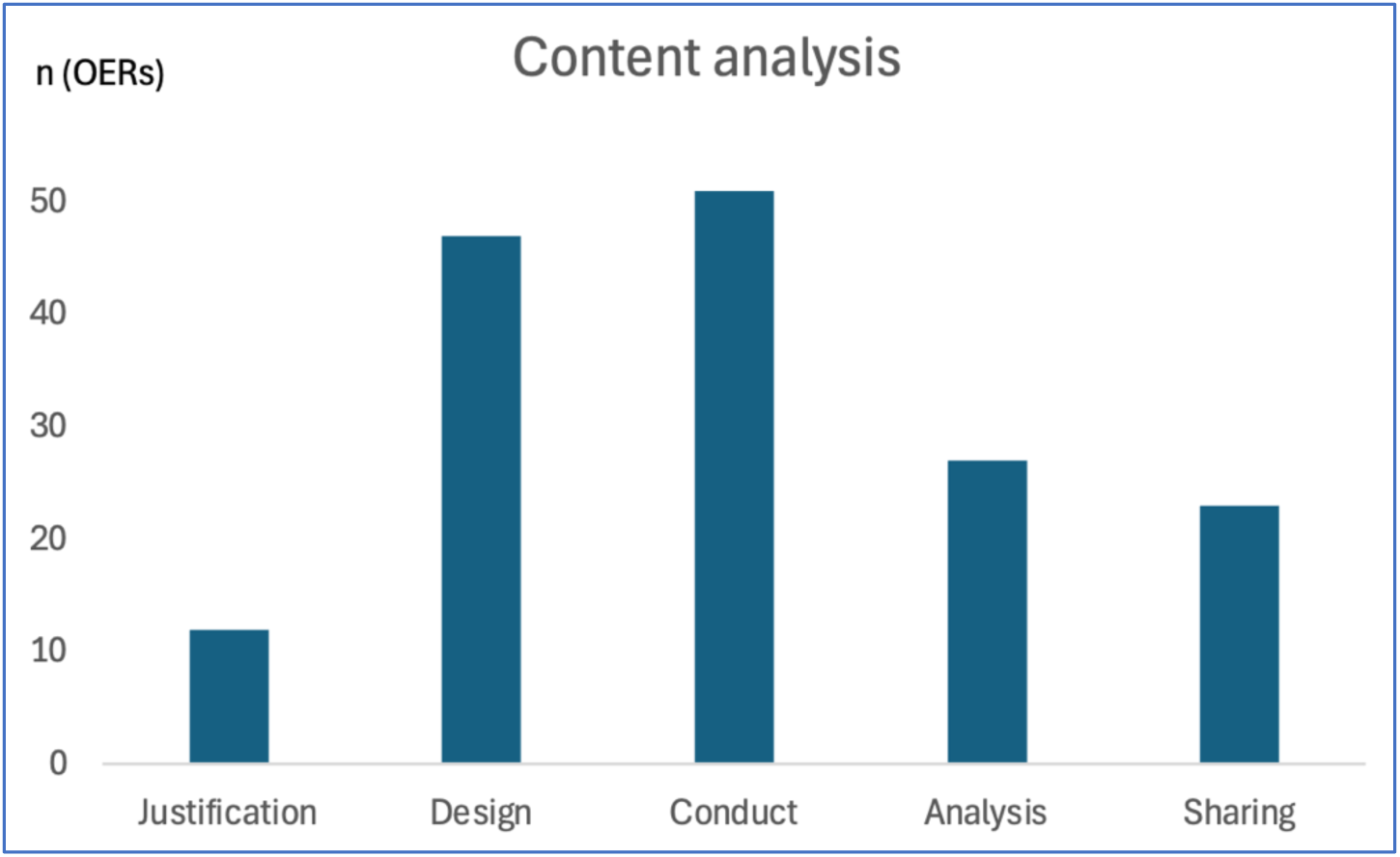
Content analysis according to trial life cycle stage. Clustering of identified open educational resource (OER) content separated into our five specified trial stages. Note that individual OERs could contain content from one or more trial stages, and that some resources contained hundreds or thousands of individual documents and tools, whereas other resources consisted of just one item. Thus, the graph is merely suggestive of the covered content.

## Discussion

### General findings

Our landscape analysis identified a large array of OERs on how to justify, design, conduct, analyse, and share RCTs. The resources varied in scope, content, quality, and their level of support by evidence. We found that most aspects of the clinical trial lifecycle were covered, yet parts of planning the trials, most importantly the justification of a new trial were rarely touched upon (Panel 4). The few resources mentioning the need for a systematic assessment of existing evidence prior to launching a new trial, such as Clinical Studies Sweden or NIHR Clinical Trials Toolkit (see ‘Planning and Design’), only did so briefly, and did not provide specific guidance or details on how to undertake this substantial task. Designated training material on how to do systematic reviews, prominently the Cochrane Handbook,^26^ also lacks guidance on how to use a systematic review to justify and design a new clinical trial. One dedicated platform, DAC from the Bill & Melinda Gates foundation, focused specifically on improving RCT informativeness, but a roadmap was absent and their best practice recommendations were not supported with references.

None of the individual resources would likely suffice as single stand-alone educational material to support the conduct of an RCT; each having some limitations or only mentioning some parts of the clinical trial life cycle superficially. Similar limitations were noted in an assessment of OERs on emergency medicine.^27^ It highlights an important aspect of OERs and a decentralised curation, which can lead to a skewedness and neglect of certain topics and over prioritisation of other topics. There are likely multiple explanations to the absence of one or more one-stop RCT resources: it is a broad and complex topic with many perspectives to cover; such resources require substantial investment and funding to develop and maintain; and entire clinical trial master degree programs exist, which might be difficult to compete with.^28, 29^

In 2022, to coordinate the global activities of clinical trials, the World Health Organization (WHO) initiated a dedicated programme of work to strengthen the clinical trials’ ecosystem.^30^ The published guidance aimed specifically to identify and propose best practices for clinical trials conduct. The WHO report summarised high-level scientific and ethical principles underlying informative clinical trials, including the importance of justifying a new clinical trial with systematic reviews.^30^

### Online learning in context

OER face several challenges compared to traditional teaching material, most importantly the lack of coordinated ‘quality control’ (i.e. peer review and/or editorial handling) and indexing (i.e. meaning that it may be invisible or hard to find in the sea that is the Internet). These limitations were highlighted already 25 years ago.^31^ However, online learning is rapidly adapting, for instance with peer reviewed medical education videos,^32^ accredited online learning material,^33^ micro credentials and adaptive learning pathways,^34^ and recommendations for making online training material findable and accessible.^35^

Like any other intervention in healthcare, the benefits of online learning about RCTs, such as easy access to information, improved user knowledge and uptake, and eventually better conducted RCTs should be adequately assessed and weighed against the potential for harms, like no user knowledge gain, wasted resources on production, wasted time for the users, and poorer RCTs.^36^ Systematic reviews of trials (conducted before the COVID-19 pandemic) comparing the effectiveness of e-learning with traditional classroom teaching of healthcare students and professionals reported ambiguous effects. A review of 59 RCTs (6750 students) found that e-learning was equivalent to, or better than, traditional teaching measured on knowledge gain and course satisfaction.^37^ A review of 16 RCTs (5679 healthcare professionals) found small to no differences measured on skills, knowledge, and on patient outcomes.^38^ Another systematic review including seven RCTs (638 healthcare students and professionals) found insufficient evidence on e-learning’s impact on patient outcomes.^39^

The comparison between blended learning and traditional teaching seems more favourable:^40^ a systematic review of postgraduate physician training (74 RCTs and 19 cluster trials; 16.895 participants) found no, or better, effects of blended learning assessed on knowledge outcomes.^41^ Another systematic review of RCTs and non-randomised studies across medical education settings (56 studies; 9.943 participants) found mainly superior knowledge outcomes of blended learning.^42^ These trials were also conducted before the COVID-19 pandemic. User attitude, uptake, and interest in online learning has likely changed as has the educators’ design, delivery, and implementation considerations.^43^ Thus, the generalisability of these results may be limited.

### Recommendations for future OER research and design

There are no agreed core outcome sets in interventional education studies to assess the effectiveness of different teaching modalities.^44^ Furthermore, the cost savings associated with online learning compared to traditional teaching may not be as favourable as one would expect.^45^ This highlights the need for further research assessing how to best design, deliver, and measure online learning experiences.^46^ It also highlights that new open educational resources must take into consideration existing evidence to be cost-effective. A 2022 survey of 7000 university students across five continents identified that most students are not content with current e-learning options and that clear course structures and roadmaps are valued the most.^47^ In another survey of 2000 students who, due to the COVID-19 pandemic, were taught a course in evidence-based medicine in three different modes (traditional classroom, online only, or blended learning), the blended approach was superior both objectively (student test scores and fail rates) and subjectively (student satisfaction).^48^ We summarise five key feature suggestions for developing future OERs on RCTs in Panel 5.

**Panel 5.**
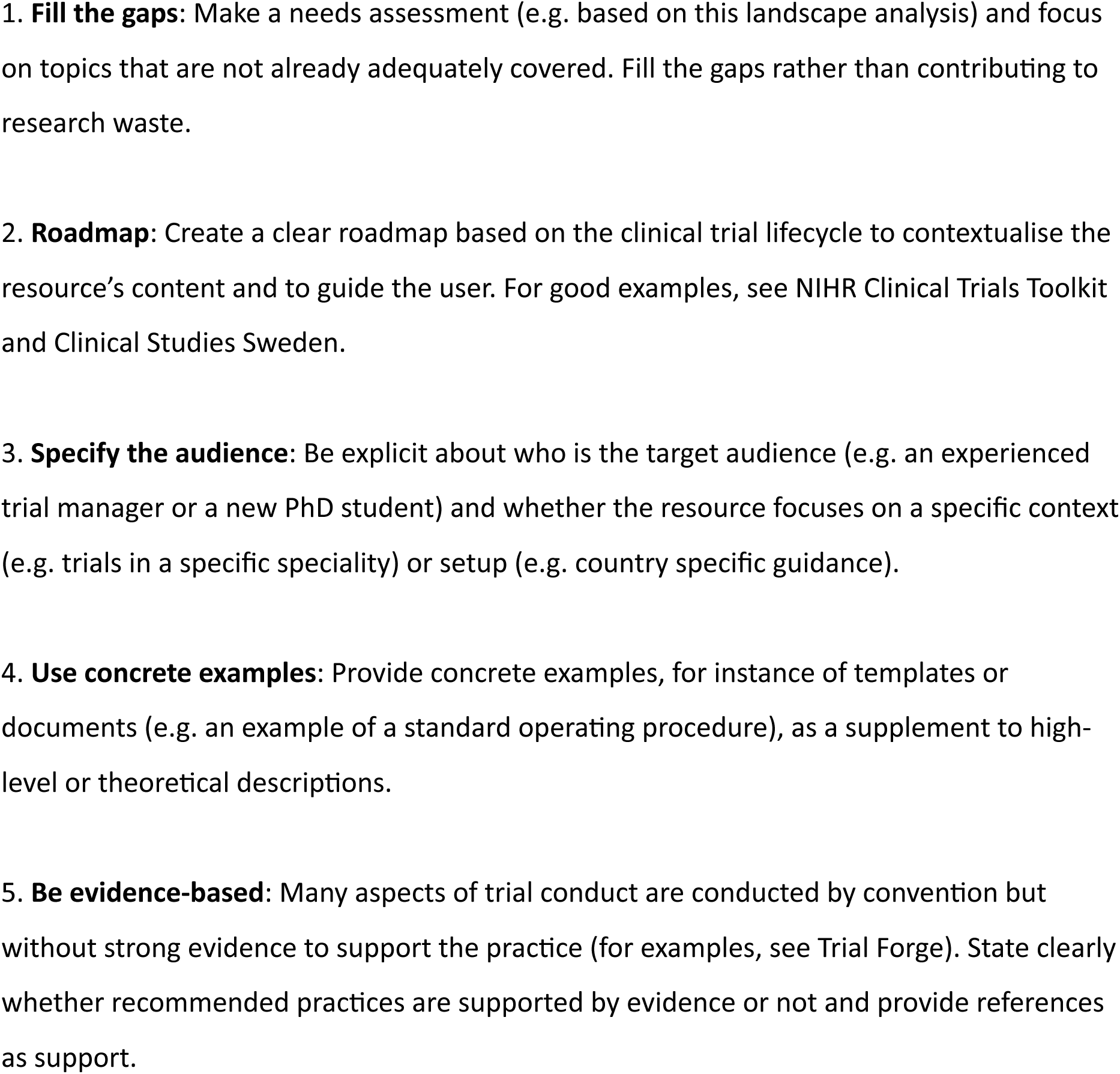
Suggestions for designing open educational resources on randomised clinical trials

### Limitations to our analysis

This landscape analysis has limitations. Firstly, our decision to use ASReview and screen only 1% of the total sample of records from our systematic search is acknowledged. We decided that the cost-benefit of screening more hits would likely yield few to none extra eligible resources based on the plateauing recall curve (Panel 3). Secondly, the topic and the significant variability in the types of resources assessed, ranging from dedicated learning platforms to individual videos, made it challenging to establish strict inclusion/exclusion criteria. This complexity led to many potentially questionable decisions, such as determining whether a specific part of a website should be included or whether a single short video qualifies as educational material. As a result, this landscape analysis does not represent an exhaustive collection of all educational resources. However, we believe to have captured the most visible (and potentially impactful) resources. Thirdly, we did not include individual published academic papers or overviews, such as this paper on web-based resources for statisticians working in the pharmaceutical industry.^49^ We searched systematically for academic papers describing OERs, whereas we did not search systematically for relevant individual papers on topics pertaining to the trial life cycle stages. Such efforts would have required a large number of systematic reviews, which was beyond the scope of this project. Fourth, we did not analyse each individual OER in minute detail, as we determined that these resources are likely frequently updated (or even removed), thus it offers limited value to most readers compared to providing a simple overview. We also abstained from in-depth quantitative analyses of the format and content, for example, if the recommended practice was evidence-based. Fifth, our content analysis is limited by the unit-of-analysis fallacy; some resources were exhaustive and contained hundreds or thousands of individual guidance, tools, and instructions while other resources were solitary guidance or videos. The content analysis is therefore only suggestive of the OER landscape. Finally, most included resources came from a few major institutions and organisations, such as NIH and NIHR, and the generalisability of material from these institutions may not be applicable for investigators in every setting. This analysis should be considered a live document that ought to be regularly updated; thus, readers are encouraged to share relevant resources that we have not identified.

## Conclusions

We identified a plethora of OERs on RCTs; however, neither individually nor collectively did they comprehensively cover all aspects of conducting justified, efficient, and informative clinical trials. Most evidently, the need for conducting updated systematic reviews as part of the planning and justification of a new clinical trial was absent from most assessed resources. Future OERs on RCTs may also consider creating instructive roadmaps to guide users and fill the gaps identified in the existing catalogue of content from this analysis with evidence-supported guidance and instructions.

## Data Availability

The complete dataset is available from the Zenodo repository at https://doi.org/10.5281/zenodo.15410961.

## Acknowledgements

KB conceived the idea and wrote the first draft. SLK did the systematic searches. All authors revised the manuscript, and all agreed on the final version. We thank Vasee Moorthy, John Owuor, and Wei Zhang from WHO’s Science Division for input on earlier drafts.

## Transparency statement

The lead author (the manuscript’s guarantor) affirms that the manuscript is an honest, accurate, and transparent account of the study being reported; that no important aspects of the study have been omitted; and that any discrepancies from the study as originally planned (and, if relevant, registered) have been explained.

