## Appendix for "Mapping open educational resources on how to justify, design, conduct, analyse, and share randomised clinical trials: a landscape analysis"

### Table of content

Supplement 1. Search strategies

Supplement 2. ASReview training

Supplement 3. Amendments compared to our protocol

Supplement 4. Full table of included resources

**Supplement 1. Search strategies**

**MEDLINE ALL Ovid (1946 to 28 February 2024)**

1. ('open access' adj3 (education* or web-site* or meducation*)).ti,ab.
2. (((online or video or digital or web-based) adj3 (resource* or material* or program* or education* or training* or course* or learning*)) or e-learning or 'blended learning' or 'micro learning').ti,ab.
3. 1 or 2

**Embase Ovid (1974 to 28 February 2024)**

1. ('open access' adj3 (education* or web-site* or meducation*)).ti,ab.
2. (((online or video or digital or web-based) adj3 (resource* or material* or program* or education* or training* or course* or learning*)) or e-learning or 'blended learning' or 'micro learning').ti,ab.
3. 1 or 2

**CINAHL (Ebsco host; 28 February 2024)**

S3 S1 OR S2

S2 TX ( (((online or video or digital or web-based) N3 (resource* or material* or program* or education* or training* or course* or learning*)) or e-learning or 'blended learning' or 'micro learning') ) OR AB ( (((online or video or digital or web-based) N3 (resource* or material* or program* or education* or training* or course* or learning*)) or e-learning or 'blended learning' or 'micro learning') )

S1 TX ( ('open access' N3 (education* or web-site* or meducation*)) ) OR AB ( ('open access' N3 (education* or web-site* or meducation*)) )

**Science Citation Index Expanded (1900 to 28 February 2024), Social Sciences Citation Index (1956 to 28 February 2024), Conference Proceedings Citation Index – Science (1990 to 28 February 2024), Conference Proceedings Citation Index – Social Science & Humanities (1990 to 28 February 2024) (Web of Science)**

#3 #2 OR #1

#2 TI=(((online or video or digital or web-based) N3 (resource* or material* or program* or education* or training* or course* or learning*)) or e-learning or 'blended learning' or 'micro learning') OR AB=(((online or video or digital or web-based) N3 (resource* or material* or program* or education* or training* or course* or learning*)) or e-learning or 'blended learning' or 'micro learning')

#1 TI=('open access' N3 (education* or web-site* or meducation*)) OR AB=('open access' N3 (education* or web-site* or meducation*))

**Supplement 2. ASReview training**

<u>Training the ASReview algorithm</u>

In our protocol, we contemplated to use all known references on open educational resources from various clinical fields and case studies of implementation of specific online e-learning programs (protocol references 21 to 38). We opted to include four seed references based on the presence of certain keywords like ‘evidence-based medicine’,[1] ‘e-learning course’,[2] ‘development of an internet platform’,[3] and ‘open access’ and ‘education’.[4] We chose five irrelevant references randomly using ASReview’s random search button.[5-9]

Furthermore, directly in ASReview, we searched for specific keywords to identify further potential seed references:

“Evidence-based” (10 hits, none relevant).
“Clinical trial” (10 hits, none relevant).
“Randomised clinical trial” (10 hits, none relevant).
“Randomized clinical trial” (10 hits, none relevant).
“Research methods” (10 hits, none relevant).

**Supplement 3. Amendments compared to our protocol**

***1. Searching additional resources***

We searched more resources than originally specified in the protocol. See our Data File for the full list of resources (https://doi.org/10.5281/zenodo.15410961).

*Clinical trial unit websites*

We searched an additional 17 US based clinical trials, in addition to the one listed in the protocol. We also searched the 14 individual ECRIN member websites.

*University or learning portals*

We searched ClassCentral, and seven more learning portals than specified in the protocol.

*Trial registries and national regulatory authorities*

These resources were not specified in the protocol. We searched the largest trial registries (ClinicalTrials.gov, the European Union Clinical Trial Register, and ISRCTN). We also searched a range of regulators, including the two largest (US Food and Drug Administration and European Medicines Agency) and a number of national regulators. We furthermore searched the European Network of Research Ethics Committees (EUREC) and the British Health Research Authority's websites.

***2. Specification of inclusion criteria for open educational resources***

The inclusion criteria were not strictly defined in our protocol due to the exploratory scope of our analysis. We decided to exclude solitary material that was not intended as educational material, e.g. protocol templates, standard operational procedures, and other available documentation, unless this was available along other documents or guidance, in which case we would classify it as 'scattered website material'.

#### ***3. Specification of how to categorise included resources***

*Categorisation of content*

In the protocol, we had not prespecified how to categorise the included resources. We decided to do this to better map which content was covered and which content was not covered. We defined five types of OER (e-learning, dedicated trial portals, scattered website material, e-books, and videos), and four formats (guidance, research publications, documentation templates, and tools).

*Definition of the five trial stages*

In the protocol, we stated to describe the curriculum of each included resource without operationalising this further. We subsequently defined five specific trial stages (justification, design, conduct, analysis, reporting) and categorised the content of each resource according to this staging.

### Supplement 4. Full tables of included resources

The 23 key resources from the main article's Panel 2 are highlighted in these five tables.

Note that the resource names are live links to the relevant websites.

#### Table 1. Dedicated portals (n=22)

| Resource | Funder | Synopsis | Content |
|---|---|---|---|
| **Australian Clinical Trial Alliance** | Public Australian funding | Dedicated platform with learning material on how to conduct clinical trials, including webinars, guidance documents, and extensive toolkits. | 1, 2, 3, 4, 5 |
| **Clinical Studies Sweden** | Swedish Regions | Dedicated portal to the conduct of RCTs with guidance, templates, and a nice roadmap | 1, 2, 3, 4, 5 |
| **Clinical Trial Information System (CTIS): online training modules** | European Union | Dedicated learning portal with 23 modules covering the CTIS reporting system and pertinent topics on trial conduct and reporting. | 2, 5 |
| **Clinical Trials Kiosk** | University of Alabama at Birmingham | Collection of guidance, templates, and tools about conducting clinical trials (focused on local infrastructure). | 3 |
| Clinical Trials Transformation Initiative | U.S. FDA and various commercial and non-commercial institutions | Dedicated portal for improving RCT conduct with guidance, research publications, and tools. | 2, 3, 5 |
| COMET | EC, MRC, and NIHR | Collection of guidance and research publications about core outcome sets. | 2 |
| **Design, Analyze, Communicate (DAC)** | Bill & Melinda Gates Foundation | Dedicated platform to make trials more informative, including best practice guidance, videos, links to external tools, and embedded e-learning courses. | 1, 2, 3, 4, 5 |
| **ECRIN Tools** | European Clinical Research Infrastructure Network | Dedicated platform with various tools for specific topics, e.g. regulatory aspects or platform trials, with guidance, publications, tools, and templates. | 2, 3 |
| **ENERI** | EU Horizon 2020 | Dedicated learning platform on research ethics with 5 e-learning modules (ENERI Classrom), a decision-tree tool, and extensive guidance documents. | 2, 3, 5 |
| **EUPATI – Open Classroom** | EU | Dedicated platform on patient involvement in clinical research with toolbox and multiple e-learning courses. | 1, 2, 3, 4, 5 |
| **Global Health Network** | Various NGOs and the University of Oxford | Learning portal with guidance, collections of links, and e-learning courses on randomised clinical trials, e.g. GCP and data monitoring safety boards. | 1, 2, 3, 5 |
| Good Trials | Good Clinical Trials Collaborative (NGO) | Dedicated website with high-level guidance to conduct clinical trials. | 2, 3 |
| NHS Health Research Authority | HRA | Dedicated website for investigators on how to plan clinical trials and submit regulatory submissions. | 1, 2, 3 |
| **NIHR Clinical Trials Toolkit** | NIHR | Dedicated platform on trial conduct with a clear roadmap, guidance and templates. | 2, 3, 4, 5 |
| **NIH Pragmatic Trials Collaboratory** | NIH | Dedicated platform for designing, conducting, and sharing clinical trials with guidance, templates, and tools. | 2, 3, 4, 5 |
| **NorCrin Method's book** | Norwegian Regions | Dedicated catalogue of Standard Operating Procedures pertaining to all steps of the trial life cycle. | 1, 2, 3 |
| **Research Methods Resource** | NIH | Dedicated platform on research methods for RCTs focusing on the design of trials with guidance, publications, and tools. Also guidance to NIH specific requirements. | 2, 3 |
| **SCTO Platforms** | Swiss Clinical Trial Organisation | Dedicated platform on clinical trials with guidance, publications, tools, and templates. | 2, 3, 4, 5 |
| TrialDesign | Private initiative (MD Anderson faculty) | Dedicated website on how to design oncology trials with guidance and publications. | 2, 3, 4 |
| **TMRN (Trials Methodology Research Network) Online Training Resource** | Health Research Board, Ireland | Dedicated platform for trial methodology with extensive training repository mainly including webinars. | 1, 2, 3, 4, 5 |
| **Trial Forge** | University of Aberdeen | Dedicated portal for improving RCT conduct (focus mainly on recruitment and retention) with guidance and research publications. | 2, 3 |

| | | | |
|---|---|---|---|
| [Training and Resources in Research Ethics Evaluation (TREEE)] | Various universities and NGOs | E-learning platform on research ethics and resources on national regulatory ethics frameworks. | 1, 2, 3 |

### Table 2. e-learning (n=20)

| Resource | Funder | Synopsis | Content |
|---|---|---|---|
| **Clinical trials operations specialization** | Johns Hopkins University | Coursera course on the conduct of RCTs, divided into four individual topic specific courses. | 2, 3, 4, 5 |
| Clinical Trials: Good Clinical Practice Specialization | Novartis | Coursera course focusing on GCP. | 3 |
| Comparative Effectiveness and Research training instruction | MD Anderson | EdX course on general RCT concepts, divided into five topic specific courses. | 2, 3, 4 |
| **Conscious II Program** | EU Erasmus Plus Programme | e-learning course in 12 chapters on full RCT conduct. | 2, 3, 4, 5 |
| Data management for clinical research | Vanderbilt University | Coursera course on data management in RCTs. | 3, 5 |
| Data sciences in pharma - patient centered outcomes research | Genentech | Coursera course on how to develop, measure and validate patient reported outcomes in RCTs. | 2 |
| **Design and interpretation of clinical trials** | Johns Hopkins University | Coursera course and general overview of RCTs. | 2, 3, 4, 5 |
| GCP Units | Danish GCP units | e-learning course on GCP (in Danish). | 3 |
| ENGAGE | AVAC (Advocacy, Access, Equity) (NGO) | e-learning course on Good Participatory Practice. | 3 |
| Faster Together | Vanderbilt University | Coursera course on involving minorities in RCTs. | 3 |
| Hands on clinical reporting using R | Genentech | Coursera course on data management and reporting of RCTs. | 4, 5 |
| Making data science work for clinical reporting | Genentech | Coursera course on data reporting of RCTs using data science. | 5 |
| NIDA training | National Institute on Drug Abuse (NIDA) | e-learning course on GCP. | 3 |
| **NIH Introduction to the Principles and Practice of Clinical Research** | NIH | e-learning course and webinar collection on how to plan, design, conduct, analyse and share randomised clinicals (accessible until 1 August 2025). | 1, 2, 3, 4, 5 |
| North Caroline Community Colleges: Introduction to Clinical Research | BioNetwork | e-learning courses on clinical trials. | 3 |
| Pragmatic and Group-Randomized Trials in Public Health and Medicine | NIH | e-learning course on pragmatic RCTs. | 2, 4 |
| STAT 509: design and analysis of clinical trials | Pennsylvania State University | Course notes about the general concepts of RCTs. | 2, 4 |
| The Simplest Guide™ to clinical data analysis with SAS | Packt | Coursera course on RCT documents and how to make clinical study reports. | 5 |
| Thinking Critically: Interpreting Randomized Clinical Trials | Stanford University | EdX course on general RCT concepts. | 2 |
| Vaccelerate Study Nurse Course | University of Cologne and EU Horizon 2020 | e-learning course on GCP for study nurses. | 3 |

### Table 3. Scattered websites (n=10)

| Resource | Funder | Synopsis | Content |
|---|---|---|---|
| Danish Medicines Agency - Guidance for non-commercial clinical trials | Danish Medicines Agency | Guidance, links to further information (on TrialNation), and templates to protocols and contracts. | 2, 3 |
| Duchenne Muscular Disease Hub Toolkit | Duchenne UK* | Collection of guidance, research publications, and documentation templates for conducting DMD trials. | 2, 3 |
| EMA scientific guidance | European Medicines Agency | Collection of guidance documents issued by EMA on various topics related to the design, conduct, and analysis of commercial pivotal trials. Also guidance related to design of pivotal disease specific trials. | 2, 3, 4 |
| Ethics and Data Protection Decision Tree | EU | GDPR decision tree to identify data protection issues during research projects, | 3 |
| FDA Guidance Documents | US Food and Drug Administration | Collection of guidance documents issued by FDA on various topics related to the design, conduct, and analysis of commercial pivotal trials. Also guidance related to design of pivotal disease specific trials. | 2, 3, 4 |
| HIV Prevention Trials Network "Manual of procedures" | Various federal US agencies | Collection of instructions and documentation templates for conducting trials in their HIV trial network. | 2, 3, 5 |
| MRC Clinical Trials Unit "Methodology" | MRC | Collection of guidance, tools and links within five domains (design, conduct, analysis, meta-analysis, software). | 2, 3, 4 |
| University of Oxford "Clinical trials and research governance" | University of Oxford | Overview of trial flow, guidance and templates (related to local infrastructure) | 2, 3 |
| Trial Methodology Research Partnership *"Guidance pack"* | MRC | Collection of guidance and publications. | 2, 3, 4, 5 |
| UKCRC *"guidance for CTUs"* | UKCRC Registered Clinical Trials Units | Collection of guidance, publications, and templates for trial units, including an e-book on monitoring trials. | 2, 3, 4, 5 |

* Not stated explicitly. Duchenne UK is a non-profit charity, unknown sponsors.

### Table 4. Videos (n=9)

| Resource | Funder | Synopsis | Content |
| --- | --- | --- | --- |
| Best Practices for Integrating Patient Reported Outcomes in Oncology Clinical Trials | National Cancer Institute | Webinar series about the development, use and analysis of patient reported outcomes in oncology trials. | 2, 4 |
| Clinical Trials Methodology Course | National Institute of Neurological Disorders and Stroke | Collection of recorded lectures from annual research courses on RCT methods. | 2, 3, 4 |
| Crash Course on Clinical Trials | Private blogger (The Brown Feminist, Canada) | Collection of videos on various topics related to clinical trials. | 3 |
| Crash Course to Clinical Research | Private blogger (Dan Sfera) | Solitary video explaining the concepts of randomised clinical trials. | 2, 3 |
| FDA Clinical Investigator Training course 2024 | FDA | Collection of recorded lectures and handouts about the general conduct of RCTs. | 2, 3, 4 |
| **GCP Mindset** | Clinical Research Organisation (GCP Service) | Collection of short videos on various topics, mainly GCP specific. | 3, 4 |
| Mind the gap: webinar series | NIH | Collection of webinars on research topics not-exclusively related to RCTs. | 2, 4 |
| NIA Clinical Trials Safety Training | National Institute of Aging | Solitary video on adverse event handling in clinical trials | 3 |
| Registering and results reporting | NIH | Solitary video on registering and results reporting on ClinicalTrials.gov | 5 |

**Table 5. e-books (n=2)**

| Resource | Funder | Synopsis | Content |
|---|---|---|---|
| Data Safety Monitoring Board Training Manual | Tufts University | e-book on data safety monitoring boards. | 3 |
| **UK Trial Managers' Network "The Guide to Efficient Trial Management"** | University of Oxford | e-book describing all steps in trial conduct, including details on local and national infrastructure | 1, 2, 3, 4, 5 |

EC = European Commission, FDA = US Food and Drug Administration, GCP = Good Clinical Practice, MRC= Medical Research Council, NGO = Non-governmental organisation, NIH = National Institutes of Health, NIHR = National Institute for Health and Care Research, RCT = Randomised clinical trial.
